# Breast and colorectal cancer awareness in Malaysians and barriers towards screening: A systematic review

**DOI:** 10.1101/2022.02.21.22271312

**Authors:** Darishiani Paramasivam, Désirée Schliemann, Maznah Dahlui, Michael Donnelly, Tin Tin Su

**Author notes:** Corresponding author: Darishiani Paramasivam /, Tin Tin Su/.

## Abstract

**Background:** Breast cancer (BC) and colorectal cancer (CRC) are considered primary cancers that affect both male and females globally. In Malaysia, BC is the most commonly diagnosed cancer among women of all ethnic groups and CRC is the second most common cancer in males and the second most common cancer in females. This systematic review was carried out to assess cancer symptom awareness and barriers to undergoing cancer screening for BC and CRC.

**Methods:** A pre-defined search was conducted between January 2008 and December 2018 using the following databases: MEDLINE, Embase, CINAHL, Web of Science, PsycINFO, Scopus and Cochrane Library for relevant articles. The search was updated in June 2020. Reviewers independently performed the data extraction and quality assessment of the included study according to the Joanna Briggs Institute assessment tools.

**Result:** 22 studies met the inclusion criteria (BC n=11; CRC n=11). Nine studies assessed symptom knowledge for BC and eight for CRC. Two studies described barriers towards cancer screening for BC and one for CRC. Four CRC studies assessed symptoms knowledge and cancer screening barriers. The most commonly reported BC symptoms were ‘painless breast lump’(27.6% - 90.8%), ‘nipple discharge’ (1.6% - 74.5%) and ‘pain in breast/ breast region’ (11.5% - 82.8%) meanwhile CRC symptoms were ‘change in bowel habits (new-onset diarrhoea or constipation)’ (28.4% - 86.6%), ‘bleeding and/or bleeding from the back passage’ (11.5% - 71.9%) and ‘weight loss’ (9.3% - 83.4%). ‘Financial issue’ (10% - 17.5%) was the most frequent blockade identified towards BC screening meanwhile ‘fear of result’ (27.6% - 32.1%) for CRC screening by Malaysians. Overall the studies carried out in Malaysia, six studies on BC symptom knowledge and one study on BC screening barrier were scored as medium study quality while four studies on CRC symptom knowledge and three studies on CRC screening barriers were scored as medium study quality.

**Conclusion:** Studies described varied and overall, limited, symptom awareness and barriers towards BC and CRC screening which likely contributes to the delayed presentation of cancers in Malaysia. There is a need for improving the awareness of BC and CRC symptoms as well as the importance of screening to encourage the early presentation of symptomatic cancer patients and down-staging of cancer.

## Introduction

The global burden of cancer is expected to increase to 22 million new cases each year by 2030, which is an increase of 75% compared to 2008 (1). In Malaysia, the total number of new cancer cases diagnosed during the period from 2012 to 2016 has increased by 11,731 cases compared to the cancer cases diagnosed during the period from 2007 to 2011 (2, 3). As reported by recent Malaysian National Cancer Registry (MNCR), the five most prevalent cancers among both males and females are breast, colorectal, trachea, bronchus and lung and lymphoma and nasopharynx cancer (3). According to the statistics on causes of death 2019, both colorectal cancer (CRC) and breast cancer (BC) were ranked as the ninth and tenth leading cause of medically certified deaths in Malaysia (4).

BC is the commonest cancer among Malaysian females (33.9%) with the highest incidence in Chinese ethnic women (Age-Standardized Incidence Rate (ASR) 40.7 cases per 100,000), followed by Indians (ASR 38.1 cases per 100,000) and Malays (ASR 31.5 cases per 100,000) (3). Almost 50% of BC cases are detected at a late stage, i.e. 47.9% of BC cases were detected at stage three and four (2). CRC is the most common cancer among males (16.9%) and the second most common cancer among females (10.7%). CRC incidence rates increased by ASR 0.2 cases in males and no change in females per 100,000 population as compared with the previous five-year period (2007–2011 report and 2012–2016) (2, 3). CRC incidence increases with age and has been reported the highest among Chinese (ASR 19.6 cases per 100,000), followed by Malays (ASR 12.2 cases per 100,000) and Indians (ASR 11.0 cases per 100,000). Over half of CRC cases (63.7%) are detected at stage three and four (3).

Numerous studies have shown that early diagnosis of cancer increases the chances of survival (5-8). Although substantial improvements have been made in cancer management, delays in diagnosis and treatment are the major deterrents for improving survival, particularly in low and middle-income countries (9). Furthermore, the lack of awareness about cancer signs, symptoms as well as negative beliefs and attitudes towards cancer and cancer screening has been associated with late presentation (10-12) which results in delayed help-seeking (13-16).

The Ministry of Health recognises the importance of cancer prevention and encourages strengthening the intervention of early detection of cancer awareness in the National Strategic Plan for Cancer Control Programme (2016-2020). In recent years, several studies reported cancer awareness and barriers to screening across Malaysia, which has built supporting evidence for improving cancer prevention strategies. The purpose of this systematic review was to identify and appraise studies that assessed the level of BC and CRC symptoms awareness and barriers towards cancer screening in Malaysians.

## Methods

This systematic review was conducted according to PRISMA guidelines. The pre-defined search protocol was registered with the International Prospective Register of Systematic Reviews (PROSPERO) (17).

### Search Strategy

The search strategy was developed a priori. Scientific literature published between January 2008 and December 2018 was searched for relevant studies by one author (DP) in the following databases: MEDLINE, Embase, CINAHL, Web of Science, PsycINFO, Scopus and Cochrane Library. Grey literature was searched in Google Scholar and reference lists of relevant studies were hand-searched. The search was updated in June 2020. Searched keywords were combined as follows: (I) knowledge; symptoms; barriers; attitude; belief; perception; screening; mammography; colonoscopy OR faecal occult blood test (FOBT); AND (II) colorectal cancer; colon cancer; colorectal tumour; colorectal carcinoma; breast cancer; breast health; breast tumour OR breast carcinoma; AND (III) Malaysia. The title screen was conducted by the first author (DP). The abstract and full-text screening were conducted according to pre-defined criteria by two independent reviewers (DP and DS or MDa).

### Study selection

Cross-sectional studies and baseline findings of intervention studies were eligible for inclusion if they meet the following criteria (I) peer-reviewed and published in the English language, (II) published in the last 10 years (1st January 2008 -1st December 2018), (III) conducted in Malaysia, (IV) included adults aged 18 years and above and (V) reported outcome measures on the level of awareness about signs and symptoms of BC and/or CRC AND/OR on the barriers towards BC and/or CRC screening.

The following types of research were excluded (I) systematic reviews & meta-analysis, reviews, qualitative studies, protocols and general reports including conference proceeding if sufficient details could not be obtained, (II) studies of Malaysians living elsewhere, healthcare professionals (e.g. Doctor, nurse, pharmacy, etc.) and patients with diagnosed cancer and/ family members of cancer patients and (III) evaluation of intervention studies and studies focusing on cancer in general and/or specific cancers other than BC and CRC.

### Data extraction

Reviewers (DP and DS, MDa or TTS) independently extracted information into a pre-defined data extraction template including information regarding the study design, population, sampling method, type of cancers, survey tool, objectives and results. Results, i.e. symptoms knowledge and barriers towards cancer screening were reported as percentages. Discrepancies between reviewers were resolved through discussion until consensus was reached.

### Quality assessment

The Joanna Briggs Institute (JBI) Critical Appraisal Tool for cross-sectional studies consisted of eight questions and was used to assess the quality of the included studies (18). The same procedure of independent reviewing as described under data extraction was followed for quality assessment. The quality of the study was assessed through a checklist by listing all eight questions with answers like ‘yes’, ‘no’, ‘unclear’ and ‘not applicable’. When each question, is answered ‘yes was scored as 1, meanwhile, if they answered wrongly (‘no’, ‘unclear’ and ‘not applicable’) was scored as 0. The scores were summed together and the total score produced for the quality of a study were accumulated (score range 0 – 8). The total score was equally divided into three categories as low (0 – 2), medium (3 – 5) and high (6 – 8).

## Results

The search yielded 1,133 records of which 19 full-texts met the eligibility criteria and were included in the systematic review. A total of 376 duplicate were discarded. Fifty-one articles were subsequently removed during the abstract screening, either because they focused on BC or CRC intervention studies, or they did not access specific symptoms or screening barriers. Hence, 19 full-text articles met the criteria and were included in the systematic review (Figure 1).

### Study characteristic

Descriptive information about the included studies is presented in Table 2. Out of ten BC studies, eight studies assessed BC symptom knowledge (19-25) and two studies assessed barriers towards mammogram screening (26, 27). Out of nine CRC studies, six studies assessed knowledge of CRC symptoms (28-33), one study assessed barriers to CRC screening (34) and two studies assessed both CRC symptom knowledge and CRC screening barriers (35, 36). Sample sizes ranged between 200 to 1,453 participants for BC studies and between 80 and 2,379 participants for CRC studies. All studies were conducted in peninsular Malaysia. Furthermore, the majority of BC and CRC studies were conducted in urban areas (n=6 BC and n=5 CRC studies), followed by sub-urban areas (n=2 BC and n=1 CRC studies) and some recruited participants from both urban and rural areas (n=2 BC and n=3 CRC studies).

### Study Quality

Study quality ranged from low to medium according to the JBI Critical Appraisal Tool (Table 1). Out of 22 studies, 14 studies fall on a medium score ranging from four to six while other 8 studies fall on a low score ranging from one to three in JBI criteria. All the 19 studies were ‘not applicable’ on ‘exposure measured validly and reliably’ and ‘objective, standard criteria used for measurement of the condition’. More than 50% of the studies did not meet the criteria of ‘the outcomes measured validly and reliably’. The studies that did not meet the criteria are all because there was no validated questionnaire tool used to gather data.

**Table 1:** Quality of the selected studies

| **ID** | **Were the criteria for inclusion in the sample clearly defined?**<br>**(Yes/No/Unclear/ Not Applicable)** | **Were the study subjects and the setting described in detail?**<br>**(Yes/No/Unclear/ Not Applicable)** | **Was the exposure measured in a valid and reliable way?**<br>**(Yes/No/Unclear/ Not Applicable)** | **Were objective, standard criteria used for measurement of the condition?**<br>**(Yes/No/Unclear/ Not Applicable)** | **Were confounding factors identified?**<br>**(Yes/No/Unclear/ Not Applicable)** | **Were strategies to deal with confounding factors stated?**<br>**(Yes/No/Unclear/ Not Applicable)** | **Were the outcomes measured in a valid and reliable way?**<br>**(Yes/No/Unclear/ Not Applicable)** | **Was appropriate statistical analysis used?**<br>**(Yes/No/Unclear/ Not Applicable)** | **Score** |
| --- | --- | --- | --- | --- | --- | --- | --- | --- | --- |
| Abdul Hadi<sup>(19)</sup> | Yes | Yes | Not Applicable | Not Applicable | Yes | Yes | Yes | Yes | 6/8 |
| Abu Samah<sup>(20)</sup> | No | Yes | Not Applicable | Not Applicable | Yes | No | Unclear | Yes | 3/8 |
| Akhtari-Zavare<sup>(37)</sup> | Yes | Yes | Not Applicable | Not Applicable | No | No | Yes | Yes | 4/8 |
| Al-Dubai<sup>(22)</sup> | Yes | Yes | Not Applicable | Not Applicable | Yes | Yes | No | Yes | 5/8 |
| Al-Naggar<sup>(34)</sup> | No | Yes | Not Applicable | Not Applicable | Yes | Yes | Unclear | Unclear | 3/8 |
| Al-Naggar<sup>(26)</sup> | Yes | Yes | Not Applicable | Not Applicable | No | No | Unclear | Yes | 3/8 |
| Dahlui<sup>(23)</sup> | No | No | Not Applicable | Not Applicable | Yes | No | Unclear | Yes | 2/8 |
| Hadi<sup>(38)</sup> | Yes | Yes | Not Applicable | Not Applicable | Yes | Yes | Yes | Yes | 6/8 |
| Harmy<sup>(28)</sup> | Yes | Yes | Not<br>Applicable | Not<br>Applicable | Yes | No | Unclear | Unclear | 3/8 |
| Hashim<sup>(33)</sup> | Yes | No | Not<br>Applicable | Not<br>Applicable | Unclear | No | Yes | Yes | 3/8 |
| Hassan<sup>(27)</sup> | Yes | Yes | Not<br>Applicable | Not<br>Applicable | Yes | Yes | Unclear | Yes | 5/8 |
| Hilmi<sup>(36)</sup> | No | Yes | Not<br>Applicable | Not<br>Applicable | Yes | Yes | Unclear | Yes | 4/8 |
| Suan<sup>(30)</sup> | No | Yes | Not<br>Applicable | Not<br>Applicable | Yes | No | Unclear | Yes | 3/8 |
| Naing<sup>(35)</sup> | Yes | Yes | Not<br>Applicable | Not<br>Applicable | Yes | No | Unclear | Yes | 4/8 |
| Parsa<sup>(24)</sup> | Yes | Yes | Not<br>Applicable | Not<br>Applicable | Yes | No | Yes | Yes | 5/8 |
| Shah<sup>(25)</sup> | Yes | No | Not<br>Applicable | Not<br>Applicable | Yes | Yes | Unclear | Yes | 4/8 |
| Su<sup>(31)</sup> | No | Yes | Not<br>Applicable | Not<br>Applicable | Yes | Yes | Yes | Yes | 5/8 |
| Yan<sup>(32)</sup> | Yes | Yes | Not<br>Applicable | Not<br>Applicable | Yes | No | Yes | Yes | 5/8 |
| Yusoff<sup>(29)</sup> | Yes | Yes | Not<br>Applicable | Not<br>Applicable | Yes | No | Unclear | Yes | 4/8 |
| Lee<sup>(71)</sup> | Yes | No | Not<br>Applicable | Not<br>Applicable | Yes | No | Unclear | No | 2/8 |
| Schliemann<sup>(72)</sup> | Yes | Yes | Not<br>Applicable | Not<br>Applicable | Yes | Yes | Yes | Yes | 6/8 |
| Sindhu<sup>(73)</sup> | Yes | Yes | Not<br>Applicable | Not<br>Applicable | Yes | No | Yes | Yes | 5/8 |

**Table 2:** Characteristic of the studies

| Authors Year | Study area | Study population | Sample size (n) | Sampling technique | Outcome assessed <sup>a</sup><br>Survey tool |
| --- | --- | --- | --- | --- | --- |
| **Breast Cancer** |  |  |  |  |  |
| Abdul Hadi<sup>(19)</sup> | Penang | Women recruited from two shopping malls | 363 | Convenient sampling | Symptoms of knowledge<br>*Tool:* Self-developed, validated questionnaire |
| Abu Samah<sup>(20)</sup> | Klang Valley & Selangor | Female students recruited from public & private universities | 842 | Multistage cluster random sampling | Symptoms of knowledge<br>*Tool:* Adapted questionnaire |
| Akhtari-Zavare<sup>(37)</sup> | Klang valley | Female undergraduate students aged 20+ recruited from four public universities | 792 | Random sampling | Symptoms of knowledge<br>*Tool:* Self-developed and validated questionnaire |
| Al-Dubai<sup>(22)</sup> | Shah Alam, Selangor | Women aged 18+ recruited from the shopping mall | 250 | Non-probability convenience sampling | Symptoms of knowledge<br>*Tool:* Self-developed and validated questionnaire |
| Dahlui<sup>(23)</sup> | Rawang | Women aged 20-60 recruited from households | 381 | Convenience sampling | Symptoms of knowledge<br>*Tool:* Self-developed questionnaire |
| Hadi<sup>(38)</sup> | Penang | Female university students recruited from one university | 200 | Convenience sampling (from 10 randomly selected faculties) | Symptoms of knowledge<br>*Tool:* Adapted and validated questionnaire |
| Parsa<sup>(24)</sup> | Six districts in Selangor | Female teachers recruited from 24 secondary schools | 425 | Multi-stage random sampling of schools | Symptoms of knowledge<br>*Tool:* Self-developed and validated questionnaire |
| Shah<sup>(25)</sup> | Kuala Lumpur | Women aged 18+ recruited from public areas (shopping malls, recreational parks, bus terminals) | 384 | Convenience sampling | Symptoms of knowledge<br>*Tool:* Adapted questionnaire |
| Lee<sup>(71)</sup> | Penang, Ipoh, Kuala Lumpur, Kota Bharu and Kota Kinabalu | Female aged 21+ recruited | 346 | Simple random sampling | Symptoms of knowledge<br>*Tool:* Modified questionnaire |
| Al-Naggar<sup>(26)</sup> | Shah Alam, Selangor | Women aged 40+ recruited from households | 200 | Random sampling of areas and apartments | Barriers towards mammogram screening<br>*Tool:* Self-developed questionnaire |
| Hassan<sup>(27)</sup> | Subang Jaya | Women aged 40+ were recruited from a private tertiary hospital | 1453 | Convenience sampling | Barrier towards mammogram screening<br>*Tool:* Self-developed and validated questionnaire |
| **Colorectal cancer** |  |  |  |  |  |
| Suan<sup>(30)</sup> | Bayan Lepas, Georgetown, Sungai Petani, Ipoh, Perak, Damansara, Petaling Jaya, Bandaraya Melaka, Johor Bahru | Males and females recruited from shopping malls | 2408 | Convenience sampling | Symptoms of knowledge<br>*Tool:* Adapted questionnaire |
| Su<sup>(31)</sup> | Perak, rural areas | Males and females aged 18+ recruited from households | 2379 | Multi-stage sampling (random and purposive sampling) | Symptoms of knowledge<br>*Tool:* Colorectal Cancer Awareness Measure questionnaire |
| Yan<sup>(32)</sup> | Serdang Hospital, Selangor | Males and females aged 18+ who accompanied relatives to the outpatient clinic | 308 | Cluster sampling | Symptoms of knowledge<br>*Tool:* Colorectal Cancer Awareness Measure questionnaire |
| Hashim<sup>(33)</sup> | Universiti Kebangsaan Malaysia Medical Centre, Selangor | Males and females aged 40+ with new onset of rectal bleeding | 80 | Not mentioned | Symptoms of knowledge<br>*Tool:* Self-developed, validated questionnaire |
| Schliemann<sup>(72)</sup> | Rawang, Selangor | Males and females aged 40+ recruited from household | 954 | Random sampling | Symptoms of knowledge<br>*Tool:* Validated questionnaire |
| Sindhu<sup>(73)</sup> | Klang Valley, Selangor | Male and females aged 18+ recruited from government health clinics | 426 | Random sampling | Symptoms of knowledge<br>*Tool:* Validated Bowel CAM questionnaire |
| Harmy<sup>(28)</sup><br>Yusoff<sup>(29)</sup> | West Malaysia | Males and females aged 50+ recruited from 44 health clinics | 1905 | Stratified multistage random sampling of clinics | Symptoms knowledge and barriers towards CRC screening<br>*Tool:* Self-developed, validated questionnaire |
| Naing<sup>(35)</sup> | Sendayan town, Negeri Sembilan | Males and females aged 18+ recruited from households | 260 | Cluster sampling of housing estates and convenience sampling of residents | Symptoms knowledge and barriers towards CRC screening<br>*Tool:* Self-developed questionnaire |
| Hilmi<sup>(36)</sup> | Kuala Lumpur | Males and females who accompanied patients to the general medical clinic in the hospital | 991 | Convenience sampling | Symptoms knowledge and barriers towards CRC screening<br>*Tool:* Self-developed tool and Champion Health Belief Model Scale |
| Al-Naggar<sup>(34)</sup> | Umra Private Hospital, Selangor | Male and female outpatients who visited the hospital | 187 | Convenience sampling | Barrier towards CRC screening<br>*Tool:* Self-developed questionnaire |

Only outcome measures of interest for this systematic review are summarised here (some studies included additional measures)
For housing areas, refer to household surveys where participants were approached who lived in the selected districts, villages or housing estates.

### Breast Cancer

#### Symptom knowledge

Awareness about 16 BC symptoms was reported in eight studies. All the symptoms of knowledge were evaluated through the cross-sectional method. Out of nine studies, three studies are low quality while the other six studies are medium quality. The most commonly surveyed BC symptoms were ‘painless breast lump’, ‘nipple discharge’ and ‘pain in breast/ breast region’ (Table 3a). The most commonly recognised symptoms were ‘painless breast lump’ (27.6% - 90.8%), ‘nipple discharge’ (1.6% - 74.5%) and ‘pain in breast/ breast region’ (11.5% - 82.8%). Furthermore, more than 50% of participants from six out of nine studies recognised ‘painless breast lump’ (58.1% - 90.8%), and seven out of nine studies recognised ‘nipple discharge’ (65.9% - 74.5%) as symptoms of breast cancer. ‘Lump under armpit’ (69.0% - 78.6%), ‘change in breast shape’ (3.4% - 81.5%) and ‘nipple retraction’ (11.5% - 82.8%) were few symptoms of breast cancer which were discussed in four studies. The least commonly reported symptoms of breast cancer were ‘painful lump’ (17.6%) (23), ‘asymmetry of the breast’ (44.8%) (22), ‘change in breast size’ (41.5%) (24), ‘breast mass’ (61.0%) (20) and ‘itchiness and discolouration of nipple’ (41.6%) (22). Across studies, more than 65% of respondents consistently identified ‘lump under armpit’ (69.0% ^-^ 78.6%) as a breast cancer symptom. Besides that, all the reviewed studies reported symptom knowledge as a percentage (%) except for one study which reported means. In this study, respondents were significantly unaware of BC non-lump symptoms such as changes in the position of the nipple, retraction of the nipple, puckering or breast dimpling, breast pain, armpit pain, a rash of the nipple, redness of the skin of the breast, change in size and shape of the breast and nipple compared to BC lump symptoms.

**Table 3:** Symptoms awareness and barriers for BC

| **Breast Cancer** | **Findings<sup>a</sup>** |
| --- | --- |
| **Signs and symptoms** |  |
| <sup>b</sup>(Painless) breast lump | 27.6% <sup>(23)</sup>, 32.2% <sup>(19)</sup>, 58.1% <sup>(25)</sup>, 61.0% <sup>(24)</sup>, 72% <sup>(38)</sup>, 73.5% <sup>(37)</sup>, 89.3% <sup>(20)</sup>, 90.8% <sup>(22)</sup> |
| Painful lump | 17.6% <sup>(23)</sup> |
| Lump under armpit | 69.0% <sup>(25)</sup>, 73.5% <sup>(19)</sup>, 78.5% <sup>(38)</sup>, 78.6% <sup>(20)</sup> |
| Swollen axillary glands | 31.4% <sup>(37)</sup>, 59.7% <sup>(20)</sup> |
| <sup>c</sup>(Bloody) nipple discharge | 1.6% <sup>(23)</sup>, 47.6% <sup>(37)</sup>, 65.9% <sup>(25)</sup>, 69.4% <sup>(24)</sup>, 70.4% <sup>(20)</sup>, 71.2% <sup>(19)</sup>, 72.1% <sup>(22)</sup>, 74.5% <sup>(38)</sup> |
| Change in breast shape | 3.4% <sup>(23)</sup>, 77.9% <sup>(25)</sup>, 78.2% <sup>(19)</sup>, 81.5% <sup>(38)</sup> |
| Asymmetry of the breast | 44.8% <sup>(22)</sup> |
| Change in breast size | 41.5% <sup>(24)</sup> |
| Discoloration of breast | 46.8% <sup>(22)</sup>, 68.5% <sup>(20)</sup> |
| Pain in breast/ breast region | 11.5% <sup>(20)</sup>, 11.5% <sup>(23)</sup>, 31.5% <sup>(24)</sup>, 78.5% <sup>(19)</sup>, 78.5% <sup>(38)</sup>, 82.8% <sup>(25)</sup> |
| Breast mass | 61.0% <sup>(20)</sup> |
| Enlargement of neighbouring lymph nodes | 75.2% <sup>(22)</sup>, 79.1% <sup>(20)</sup> |
| Dimpling of breast skin | 50.8% <sup>(25)</sup>, 58.5% <sup>(20)</sup>, 60.3% <sup>(19)</sup> |
| Breast skin retraction | 48.4% <sup>(22)</sup>, 66.2% <sup>(20)</sup> |
| Nipple retraction | 16.6% <sup>(24)</sup>, 25.8% <sup>(37)</sup>, 46.8% <sup>(22)</sup>, 74.5% <sup>(20)</sup> |
| Itchiness and discolouration of nipple | 41.6% <sup>(22)</sup> |
| **Barriers toward screening** |  |
| Lack of time | 42.5% <sup>(26)</sup> |
| Do not know the purpose of mammogram screening | 32% <sup>(26)</sup> |
| Don't know where to go for the test | 53.5% <sup>(26)</sup> |
| Afraid of the test result | 20% <sup>(26)</sup> |
| Costly | 10 % <sup>(27)</sup>, 17.5% <sup>(26)</sup> |
| Unnecessary test | 14% <sup>(26)</sup> |
| The perception that they are not at risk | 30 % <sup>(27)</sup> |
| The perception that mammogram screening is painful | 20 % <sup>(27)</sup> |
| The perception that breast cancer treatment is painful | 54.9 % <sup>(25)</sup> |

<sup>a</sup> Proportion of participants (%) that reported knowledge/ awareness about the following symptoms / reported the following barriers towards breast cancer screening
<sup>b</sup> Some studies specified as painless lump whereas other studies only specified as a breast lump
<sup>c</sup> Some studies specified as nipple discharges whereas other studies only specified as a blood discharge from nipple

#### Barriers towards screening

Only two studies explored the barriers towards BC screening in Malaysian women (26, 27) (Table 3). In those two studies, nine barriers towards mammogram screening were reported. All the barriers towards mammogram screening studies were evaluated through a cross-sectional method. Out of two studies, one was low quality while other was medium quality. Both studies assessed whether ‘financial issues’ (10% - 17.5%) was a barrier towards screening, otherwise, the two studies assessed different barriers. ‘Lack of time’ (42.5%) was the most commonly reported barrier (26), followed by ‘do not know the purpose of mammogram’ (32%) (26) and the ‘perception that they are not at risk’ (30%) (27). Furthermore, barriers such as ‘mammogram screening are painful’ (20%), ‘mammogram is an unnecessary test’ (14%) and ‘afraid of the test result’ (20%) were less commonly reported barriers.

### Colorectal Cancer

#### Symptom Knowledge

Awareness about 14 CRC symptoms was assessed in 10 out of 11 CRC studies. All the 10 studies were analysed by the cross-sectional method. Quality of seven studies was scored under medium quality while the other three studies scored under low quality. The most-reported CRC symptoms were ‘change in bowel habits (new-onset diarrhoea or constipation)’, ‘bleeding and/or bleeding from the back passage’ and ‘weight loss’ (Table 4). The most commonly recognised symptoms were ‘blood in stool’ (40.6% - 86.9%), ‘change in bowel habits (new-onset diarrhoea or constipation)’ (28.4% - 86.6%), abdominal pain (31.4% - 85.6%) and ‘weight loss’ (9.3% - 83.4%). Out of 13 CRC symptoms, 10 symptoms were recognized by >50% of participants from four out of eight studies, except for ‘piles’ (49.2%), ‘mucus in stool’ (2.2%), ‘difficulties swallowing’ (35.4%) and ‘tiredness/anaemia’ (27.7% - 35.1%), which were recognised by <50% of participants. Furthermore, the least commonly surveyed symptoms of CRC were ‘piles’ (49.2%), ‘constipation’ (53.4%), ‘mucus in stool’ (2.2%), ‘difficulties swallowing’ (35.4%) and ‘loss of appetite’ (63.5%).

**Table 4:** Symptoms awareness and barriers for CRC

| **Colorectal Cancer** |  |
| --- | --- |
| **Signs and symptoms** |  |
| Piles | 49.2%<sup>(28)</sup> |
| Constipation | 53.4%<sup>(28)</sup> |
| Change in bowel habit (new onset diarrhea or constipation) | 28.4%<sup>(31)</sup>, 32.9%<sup>(73)</sup>, 37.5%<sup>(36)</sup>, 45.9%<sup>(72)</sup>, 50.3%<sup>(32)</sup>, 67.3%<sup>(35)</sup>, 86.6%<sup>(30)</sup>, , |
| Incomplete emptying of bowel | 2.1%<sup>(73)</sup>, 26.7%<sup>(31)</sup>, 31.2%<sup>(32)</sup>, 66.8%<sup>(28)</sup>, 45.2%<sup>(72)</sup>, |
| Blood in the stool | 33.3%<sup>(73)</sup>, 40.6%<sup>(31)</sup>, 56.8%<sup>(32)</sup>, 86.9%<sup>(30)</sup>, 54%<sup>(72)</sup>, |
| Bleeding and/or Bleeding from back passage | 11.5%<sup>(73)</sup>, 34.3%<sup>(36)</sup>, 37%<sup>(31)</sup>, 46.4%<sup>(72)</sup>, 63.4%<sup>(35)</sup>, 71.1%<sup>(32)</sup>, 71.9%<sup>(28)</sup>, |
| Back passage pain | 24.6%<sup>(31)</sup>, 35.1%<sup>(72)</sup>, 3.3%<sup>(73)</sup>, 52.3%<sup>(32)</sup> |
| Mucus in stool | 2.2%<sup>(36)</sup> |
| Difficulties swallowing | 35.4%<sup>(28)</sup> |
| Lump in abdomen | 2.3%<sup>(73)</sup>, 35.5%<sup>(31)</sup>, 51.0%<sup>(32)</sup>, 74.4%<sup>(28)</sup>, 49.0%<sup>(72)</sup>, |
| Abdominal pain | 31.4%<sup>(36)</sup>, 36.1%<sup>(31)</sup>, 38.5%<sup>(73)</sup>, 51%<sup>(72)</sup>, 58.1%<sup>(32)</sup>, 85.6%<sup>(30)</sup> |
| Loss of appetite | 63.5%<sup>(28)</sup> |
| Weight loss | 9.3%<sup>(36)</sup>, 23.7%<sup>(73)</sup>, 33.1%<sup>(31)</sup>, 50%<sup>(32)</sup>, 51.8%<sup>(72)</sup>, 64.8%<sup>(28)</sup>, 83.4%<sup>(30)</sup>, |
| Tiredness/Anaemia | 14.6%<sup>(73)</sup>, 27.7%<sup>(31)</sup>, 35.1%<sup>(32)</sup>, 38.8%<sup>(72)</sup>, |
| **Barriers toward screening** |  |
| I do not know if should have faecal occult blood test (FOBT) | 50.8%<sup>(34)</sup> |
| FOBT is not necessary | 46.5%<sup>(34)</sup> |
| FOBT was not recommended by doctor | 11.2%<sup>(29)</sup>, 35.3%<sup>(34)</sup> |
| I did not do FOBT because I do not have a health problem | 40.6%<sup>(34)</sup> |
| <sup>b</sup>FOBT is a painful test | 51.5%<sup>(28)</sup>, 53.5%<sup>(34)</sup>, |
| <sup>b</sup>FOBT embarrassing | 61.5%<sup>(28)</sup> |
| <sup>b</sup>FOBT cause side effect | 54.6%<sup>(28)</sup> |
| <sup>b</sup>FOBT troublesome | 54.6%<sup>(28)</sup> |
| <sup>b</sup>FOBT expensive | 53.1%<sup>(28)</sup> |
| <sup>b</sup>FOBT uncomfortable | 51.9%<sup>(28)</sup> |
| <sup>b</sup>FOBT fear of the result | 54.3%<sup>(28)</sup> |
| <sup>b</sup>FOBT time consuming | 45.7%<sup>(28)</sup> |
| <sup>b</sup>Colonoscopy embarrassing | 67.7%<sup>(28)</sup> |
| <sup>b</sup>Colonoscopy cause side effect | 48.8%<sup>(28)</sup> |
| <sup>b</sup>Colonoscopy time consuming | 34.7%<sup>(28)</sup> |
| <sup>b</sup>Colonoscopy expensive | 35.4%<sup>(28)</sup> |
| <sup>b</sup>Colonoscopy uncomfortable | 29.1%<sup>(28)</sup> |
| <sup>b</sup>Colonoscopy painful | 46.5%<sup>(28)</sup> |
| I do not think that flexible sigmoidoscopy is necessary | 41.2%<sup>(34)</sup> |
| I have no symptoms of colorectal cancer | 12.8%<sup>(29)</sup>, 39.0%<sup>(34)</sup> |
| Screening is embarrassing | 35.2%<sup>(29)</sup>, 55.1%<sup>(34)</sup> |
| Fear of result | 27.6% <sup>(35)</sup>, 29.8% <sup>(29)</sup>, 32.1% <sup>(34)</sup> |
| Fear of discomfort | 30% <sup>(29)</sup>, 63.8% <sup>(35)</sup> |
| Fear of side effects | 41.4% <sup>(35)</sup> |
| Expensive | 22.6% <sup>(29)</sup>, 53.4% <sup>(35)</sup> |
| Not bothered | 31.9% <sup>(29)</sup> |
| Busy | 33.4% <sup>(29)</sup> |
| Time consuming | 21.8% <sup>(29)</sup> |
| Do not understand the procedure | 18.5% <sup>(29)</sup> |
| Do not know how to go about screening | 17.1% <sup>(29)</sup> |
| Troublesome | 29.4% <sup>(29)</sup> |

<sup>a</sup> Proportion of participants (%) that reported knowledge/ awareness about the following symptoms / reported the following barriers towards breast or colorectal cancer screening
<sup>b</sup> Some studies specified as barriers towards FOBT and Colonoscopy whereas others did not specify the screening method

#### Barriers towards screening

Out of the eleven CRC studies, five studies reported barriers towards CRC screening (Table 4) (28, 29, 34-36). A total of 18 barriers were assessed through the cross-sectional method from all the 5 studies. Three studies were identified with medium quality while the other two studies were with low quality. The most commonly observed barrier towards CRC screening was ‘fear of result’ (27.6% ^-^ 32.1%). The most commonly expressed barriers towards CRC screening were ‘don’t know if I should have faecal occult blood test (FOBT)’ (50.8%), ‘FOBT is a painful test’ (51.5% - 53.5%) (28, 34), ‘screening is embarrassing’ (35.2% - 55.1%), ‘fear of discomfort’ (30% - 63.8%^)^ and ‘expensive’ (22.6% - 53.4%).

## Discussion

This systematic review assessed the level of knowledge of BC and CRC symptoms and the barriers towards screening for both cancers in Malaysia. The review summarised studies conducted over the past 10 years and reviewed by cancer type, study location, cancer symptoms, and barriers to cancer screening. Reported awareness varied to a large extends between studies and was overall low despite BC and CRC being the commonest cancers in the country. Although the study included a relatively small number of studies on knowledge of BC and CRC symptoms as well as cancer screening barriers among Malaysians, more symptom knowledge studies were conducted compared to barriers towards cancer screening studies.

### Breast Cancer

#### Symptom Knowledge

Among ten studies on breast cancer, knowledge for symptoms was discussed in eight studies. All the studies were conducted in Kuala Lumpur (1), Selangor (5), and Penang (2), the most developed states in Malaysia. Level of awareness of some BC symptoms such as ‘painless breast lump’, ‘lump under armpit’, ‘swollen axillary glands’, ‘nipple discharge’, ‘change in breast shape’, ‘discolouration of the breast’, ‘pain in breast/ breast region’, ‘enlargement of neighbouring lymph nodes’, ‘dimpling of breast skin’, ‘breast skin retraction’ and ‘nipple retraction’ are consistently discussed in different locations in Malaysia, besides there are also other specific symptoms of BC that many other participants were unable to identify. ‘Painless breast lump’ and ‘nipple discharge’ were the most (19, 20, 22-25, 37, 38) and ‘pain in breast/ breast region’ (19, 20, 23-25, 38) was the second most recognised BC symptom identified in this systematic review. Of these, six studies were assessed as being medium quality. More than half of the studies showed that these were the most frequently recognised BC symptoms. The ‘painless breast lump’ and ‘nipple discharge’ were recognised as early BC symptoms by over 55% of participants from a mix of urban and suburban areas in Kuala Lumpur (1), Selangor (4), and Penang (1). However, ‘pain in breast/ breast region’ was only identified by participants from Kuala Lumpur (82.8%) and Penang (78.5%) recognized. ‘Nipple discharge’ and ‘pain in breast/ breast region’ were two symptoms that were consistent with the outcomes of research focused primarily on women from the United Kingdom (71.9% and 53.4%) (12), China (55.2% and 51.7%) (39) and Iran (86.6% and 75.6%) (40). In this systematic review, participants from selected urban and suburban areas (Malaysia) exhibited the least knowledge of ‘painless breast lump’ (27.6%), ‘painful lump’ (17.6%), ‘nipple discharge’ (1.6%), ‘change in breast shape’ (3.4%) and ‘pain in the breast/breast region’ (11.5%) as the symptoms of BC (23). This is similar to a study conducted among women from suburban area in India reported that ‘painless lump’ (13%), ‘breast pain’ (16%) and ‘nipple discharge’ (8%) except ‘painless breast lump’ (44%) as symptoms of BC (41). Besides, findings were consistent with a study which was conducted among participants from a rural area (located in south-western Nigeria) who indicated that ‘painless lump’ (1.9%) and nipple discharge (0.5%) as symptoms of BC (42). This suggests that women from rural as well as suburban areas have poor knowledge of the symptoms of BC. Poor knowledge in recognizing BC symptoms is likely to delay the seeking of health care. Moreover, the finding indicates that only five studies have addressed the ‘lump under armpit’ which is a key symptom of BC.

Consistently, more than 65% of Malaysians women recognised this as a symptom of BC. These findings were marginally higher with previous research with women from the United Kingdom, which has exhibited that ‘lump under armpit’ (86.1%) was likely to be attributed to BC (12). Besides, Grunfeld et al. reported that 80% of women surveyed from the United Kingdom recognised ‘lump under armpit’ as a symptom of BC (43). Our result indicates that three studies which were conducted among women from a mix of urban and suburban areas (Shah Alam and Klang Valley) showed that most of them are able to identify ‘breast lump’ (90.8%, 89.3% and 73.5%) compared to ‘nipple discharge’ (72.1%, 70.4% and 47.6%) as a symptom of BC. A previous study in the UK also showed similar finding of high knowledge about ‘breast lump’ (93.3%) as a symptom of breast cancer and low knowledge about ‘nipple discharge’ (71.9%) (12). In addition, three among four studies have reported that less than 50% of Malaysian women recognise ‘nipple retraction’ as a BC symptom. Two out of three studies are carried out among urban and educated women such as from Selangor living undergraduate students and school teachers. However, this finding contradicts with the study which was carried out among university students from Muscat where more than half of the them were considered above as a symptom of BC (44). This may reflect discrepancies in study designs, modes of recruitment or sampling procedures of the study.

#### Barriers towards cancer screening

The two studies that reported barriers towards BC screening were conducted among 40-year-old women from Selangor. Out of two studies, barriers towards BC screening from one study were prompted through the questionnaire, whereas the barriers were identified by the participants in another study. In this review, the most common barriers towards BC screening ‘Cost’ known the common and only less than 20% of participants are facing those barriers to BC screening. It is because BC screening (mammogram) can be carried out in 16 private clinics and hospitals registered with the National Population and Family Development Board (2007). The cost is subsidized by the Development Ministry of Women, Family and Community where it offers an RM50 subsidy for each mammography session. The finding was marginally consistent with findings of the past study by Wood and his team, which 35% of people from Santiago, Chile still facing the financial barrier (‘cost’) (45). Moreover, the results of a study conducted in Hong Kong by Margaret Chua and his team among Chinese women showed that ‘cost’ was the most frequently reported barrier to screen for BC. (46). Furthermore, slightly more than half of Malaysians still do not know ‘where to go for BC screening’ (26). This shows that there is still a lack of knowledge and awareness among the Malaysian population on the hospitals or clinics that provide BC screening service. ‘Lack of time or being too busy’ to schedule mammography was another reason mentioned by Malaysian women in this review and were assessed as low quality (26). A possible reason for the above-mentioned barriers of mammography among Malaysian women may be being reluctant about the seriousness of BC. The present review finding is in line with women aged more than 40 years from West Bank whom also mentioned that ‘lack of time or being too busy’ (47%) as the most common barrier (47). In addition, a study among low-income Asian-American participants also has reported that ‘lack of time or being too busy’ as the most common barrier (48). This was also one of the most common reasons mentioned by Chinese women in Hong Kong for reluctance to participate in mammography screening (49). This study also reported that 20% of Malaysian women are still ‘afraid of the mammogram result’ (26) and ‘believe that mammogram screening is painful’ (27). The present finding also supports Garbers and his team’s research study which concluded that low-income women from New York are ‘afraid of the mammogram result’ (50). Previous studies among Arab and Filipino women living in the United States have reported similar findings on the ‘afraid of the mammogram result’ as a barrier (51-53). Afraid of discovering cancer is more associated with the cultural impact as the dynamics of the family and the assumption of cancer fatalism will have an impact on a women’s life. This barrier could be improved by effectively addressed through culturally appropriate awareness and intervention programs. In addition, the barrier ‘believes that mammogram screening is painful’ found in this review in accordance with the barrier that stated by both Filipino and Asian-Indian women living in the United States (53). Another research found that as compared to non-obese, obese women were significantly more likely to feel ‘pain’ from mammograms (54). Other research has also shown that up to 35% of women undergoing mammograms complain of ‘pain’ (20% in this study), and that pain acts as a barrier to other women returning for mammograms screening (55). Similar findings on the above mentioned barrier to mammogram were previously been published through few studies (50, 52). By providing information on privacy and gentle handling of the breasts during the procedure can help to minimise the pain or uncomfortable feelings during BC screening among Malaysian women.

### Colorectal Cancer

#### Symptom Knowledge

Among nine studies on CRC, knowledge of symptoms was discussed in eight studies. All these studies were conducted in seven different states and those places are known as the most developed states in Malaysia. In this systematic review, a study which was conducted among friends and relatives accompanying patients to the general medical clinic in the University of Malaya Medical Centre, Kuala Lumpur were demonstrated the least (less than 40%) frequently identified unprompted symptoms of CRC such as ‘change in bowel habit’ (37.5%), ‘bleeding and/or bleeding from back passage’ (34.3%), ‘mucus in the stool’ (2.2%), ‘abdominal pain’ (31.4%), and ‘weight loss’ (9.3%). Moreover, another study among participants from the rural area of Perak (Malaysia) also showed that less than 41% of them recognized prompted symptoms of CRC correctly including ‘change in bowel habit’ (28.4%), ‘incomplete emptying of bowel’ (26.7%), ‘blood in the stool’ (40.6%), ‘bleeding and/or bleeding from back passage’ (37%), ‘back passage pain’ (24.6%), ‘abdominal pain’ (36.1%), ‘weight loss’ (33.1%) and ‘tiredness/anaemia’ (27.7%). This shows that rural as well as urban residents are unaware of CRC warning symptoms. Similarly, a study conducted among 350 outpatients of Mayo General Hospital, Ireland concluded that 26.6% of the participants were able to name at least a CRC symptom (59), suggesting that awareness is limited in low- and middle- as well as high-income countries. The outcomes of this study suggest that ‘change in bowel habits’ (30-32, 35, 36) and ‘weight loss’ (28, 30-32, 36) are the most commonly reported (or assessed??) symptoms of CRC. Less than 40% of participants from two (31, 36) studies could identify the above-mentioned symptoms as the key symptoms of CRC. The results are consistent with the results of earlier studies that conducted in the United Kingdom, which suggested that <30% of respondents recognised ‘change in bowel habits’ (11, 56-58) and ‘weight loss’ (11, 57-61) as a symptom of CRC. This also indicated that compared to participants who recall symptoms, the percentage of participants who recognized those symptoms as the key symptom of CRC was greater (31, 36). The next widely discussed symptoms were ‘abdominal pain’ (30-32, 36) followed by ‘incomplete emptying of bowel’ (28, 31, 32), ‘blood in the stool’ (30-32) and ‘bleeding’ (28, 35, 36). Out of four studies, two studies showed that more than 50% of Malaysians were able to identify ‘abdominal pain’ as one of the main symptoms of CRC, whereas two other studies show less than 50% of Malaysians. Past studies conducted among the population of Western Australia, the UK and Kuwait (57, 62, 63) indicated that more than 55% of participants were able to recognize ‘abdominal pain’ as a symptom of CRC, while another study in the UK disputes this result (58). More than 55% of Malaysians can recognize both ‘blood in the stool’ and ‘bleeding’ as CRC symptoms. Findings from this systematic review suggest that awareness about ‘blood in the stool’ and ‘bleeding’ is higher in Malaysians compared to the United Kingdom, Ireland, Gulf and Britain, which indicated that less than 40% of people were able to recognise the above symptoms (11, 56, 58-60).

#### Barriers towards cancer screening

All these studies were conducted in urban area such as Negeri Sembilan, Kuala Lumpur and Selangor among participants aged more than 18 years. In this systematic analysis, a study conducted among participants from western Malaysia was rated as low quality, but most barriers were addressed and least frequently (less than 40%) prompted barriers to CRC screening were found. This result is slightly similar with certain barriers that discussed in a recent study which was conducted among more than 50 years old population from the rural area of the USA such as ‘fear’ (25.4%), ‘financial barrier’ (25.4%), ‘no recommendation/referral’ (33.1%) and ‘discomfort’ (10.2%) (64). To overcome Malaysia’s financial barrier towards CRC screening by developing an intervention programme that focuses primarily on ways to minimize the cost of screening tests or enhance insurance coverage. Additionally, it also will be more beneficial for Malaysian to make more initiatives to improve insurance coverage and to advocate information relating to increased insurance coverage. This review also suggests that participants were prompted by researchers from two studies that ‘FOBT is a painful test’ (53.5 %) as a screening barrier, which suggests that participants do not know what an FOBT test as a providing a stool sample is unlikely to be painful. The most perceived barrier to screening was anxiety about knowing the outcome of screening (‘fear of the results’). This barrier to CRC screening was encountered by less than 35% of Malaysians. This is evidenced by a study conducted between the church members of the Appalachian area between 2002 and 2003. Less than 10% of respondents said that they were afraid to find out about the test result as the main concern for not being screened (65). Malaysian’s most recognised barriers to CRC was ‘screening is embarrassing’ (55.1%) and ‘fear of discomfort’ (63.8%). One study examined the association of embarrassment with CRC screening and found that 49% of African – Americans felt that the test was embarrassing (66). Although, another study showed that less than 10% of the Appalachians admitted that they felt uncomfortable or embarrassed to carry out a CRC screening test which was contrary to the Malaysians (65). Apart from that, slightly more than 50% of Malaysians said they did not know if they should have an FOBT (50.8%) (34) and FOBT is a very expensive test (53.4%) (35). The primary reason for Malaysia does not understand the importance of CRC screening is due to the lack of referral from the healthcare professionals and some even said that screening was not necessary. This indicates that a healthcare professional plays an important role in health decision-making for Malaysians. A study in Hong Kong identified that 86.0% of participants have agreed that cost as the main barrier to screening(67). A cross-sectional study of barriers between groups of people at risk for CRC in the Omaha Midwestern Metropolitan Area described that ‘fear of cancer diagnosis’ (42%), ‘embarrassment’ (35%) and ‘screening test costs’ (44 %) (68) were the main barriers towards screening. Poor awareness and negative perception towards CRC screening could be one of the main factors for these negative attitudes such as embarrassment, fear and pain. In particular, Leung and his team indicated that the feeling of embarrassment may be due to cultural beliefs, such as faecal aversion (69). Further, there were 83 research studies by high-income countries on the most commonly mentioned barriers to CRC screening was included in a recent systematic review. Few of them were consistent with the findings of this survey (70).

## Conclusion

Everyone over 40 years old should be educated on symptom awareness and the importance of cancer screening for both BC and CRC, which would help to lower the mortality from both BC and CRC. There is an urgent call for cancer awareness and screening programs at a national and regional level, including partnerships with community organizations and the health system. Current findings suggest there is a litter understanding as well as knowledge on early symptoms of BC and CRC, but there is a significant shortage, particularly of certain BC and CRC symptoms which would allow early detection of cancer. The health education programme should be given to the public by trained healthcare professionals to overcome those cancer screening barriers which lead to increases the uptake of cancer screening. Education on cancer awareness programs should be enhanced to spread symptoms awareness of breast as well as colorectal cancer and promote the importance of cancer screening to increase the screening uptake of both cancers.

## Data Availability

All data produced in the present work are contained in the manuscript

## REFERENCES

1. Bray F, Ren J-S, Masuyer E, Ferlay J. Global estimates of cancer prevalence for 27 sites in the adult population in 2008. International Journal of Cancer. 2013;132(5):1133–45.

2. Zainal Ariffin O, Nor Saleha IT. National cancer Registry report 2007 Ministry of Health, Malaysia2011.

3. Azizah AMHB, Nirmal K., Siti Zubaidah AR., Puteri NA., Nabihah A., Sukumaran R., Balqis B., Nadia SMR., Sharifah SSS., Rahayu O., Nur Alham O and Azlina AA. Malaysian National Cancer Registry 2012-20162019. 116 p.

4. Malaysia DoS. Statistics on Causes of Death Malaysia, 2019: Department of Statistics Malaysia; 2019.

5. Simon AE, Waller J, Robb K, Wardle J. Patient delay in presentation of possible cancer symptoms: the contribution of knowledge and attitudes in a population sample from the United kingdom. Cancer epidemiology, biomarkers & prevention : a publication of the American Association for Cancer Research, cosponsored by the American Society of Preventive Oncology. 2010;19(9):2272–7.

6. UK CR. National Awareness and Early Diagnosis Initiative (NAEDI). 2011 [Available from: http://www.cancerresearchuk.org/health-professional/early-diagnosis-activities/national-awareness-and-early-diagnosis-initiative-naedi.

7. Ironmonger L, Ohuma E, Ormiston-Smith N, Gildea C, Thomson CS, Peake MD. An evaluation of the impact of large-scale interventions to raise public awareness of a lung cancer symptom. British Journal of Cancer. 2015;112(1):207–16.

8. Konfortion J, Jack RH, Davies EA. Coverage of common cancer types in UK national newspapers: a content analysis. BMJ Open. 2014;4(7):e004677.

9. Shah SC, Kayamba V, Peek RM, Jr., Heimburger D. Cancer Control in Low- and Middle-Income Countries: Is It Time to Consider Screening? J Glob Oncol. 2019;5:1–8.

10. Brunswick N, Wardle J, Jarvis MJ. Public awareness of warning signs for cancer in Britain. Cancer Causes & Control. 2001;12(1):33–7.

11. Robb K, Stubbings S, Ramirez A, Macleod U, Austoker J, Waller J, et al. Public awareness of cancer in Britain: a population-based survey of adults. British Journal of Cancer. 2009;101(2):S18–S23.

12. Linsell L, Burgess CC, Ramirez AJ. Breast cancer awareness among older women. British Journal of Cancer. 2008;99(8):1221–5.

13. Davis TC, Rademaker A, Bailey SC, Platt D, Esparza J, Wolf MS, et al. Contrasts in Rural and Urban Barriers to Colorectal Cancer Screening. American Journal of Health Behavior. 2013;37(3):289–98.

14. Al-Azri M, Al-Maskari A, Al-Matroushi S, Al-Awisi H, Davidson R, Panchatcharam SM, et al. Awareness of Cancer Symptoms and Barriers to Seeking Medical Help Among Adult People Attending Primary Care Settings in Oman. Health Serv Res Manag Epidemiol. 2016;3:2333392816673290-.

15. Cassim S, Chepulis L, Keenan R, Kidd J, Firth M, Lawrenson R. Patient and carer perceived barriers to early presentation and diagnosis of lung cancer: a systematic review. BMC Cancer. 2019;19(1):25.

16. Gede N, Reményi Kiss D, Kiss I. Colorectal cancer and screening awareness and sources of information in the Hungarian population. BMC Family Practice. 2018;19(1):106.

17. National Institute of Health Research: PROSPERO 2018 [Available from: https://www.crd.york.ac.uk/PROSPERO/display_record.php?RecordID=94793.

18. Moola S MZ, Tufanaru C, Aromataris E, Sears K, Sfetcu R, Currie M, Qureshi R, Mattis P,, Lisy K MP-F. Chapter 7: Systematic reviews of etiology and risk. Aromataris E MZ, editor. Joanna Briggs Institute Reviewer’s Manual. The Joanna Briggs Institute2017.

19. Hadi MA, Hassali MA, Shafie AA, Awaisu A. Knowledge and Perception of Breast Cancer among Women of Various Ethnic Groups in the State of Penang: A Cross-Sectional Survey. Medical Principles and Practice. 2010;19(1):61–7.

20. Abu Samah A, Ahmadian M, Latiff LA. Insufficient Knowledge of Breast Cancer Risk Factors Among Malaysian Female University Students. Global Journal Of Health Science. 2015;8(1):277–85.

21. Akhtari-Zavare M, Juni MH, Ismail IZ, Said SM, Latiff LA. Health Beliefs and Breast Self-Examination among Undergraduate Female Students in Public Universities in Klang Valley, Malaysia. Asian Pacific Journal Of Cancer Prevention: APJCP. 2015;16(9):4019–23.

22. Al-Dubai SAR, Qureshi AM, Saif-Ali R, Ganasegeran K, Alwan MR, Hadi JIS. Awareness and Knowledge of Breast Cancer and Mammography among a Group of Malaysian Women in Shah Alam. Asian Pacific Journal of Cancer Prevention. 2011;12(10):2531–8.

23. Dahlui M, Gan DEH, Taib NA, Pritam R, Lim J. Predictors of Breast Cancer Screening Uptake: A Pre Intervention Community Survey in Malaysia. Asian Pacific Journal of Cancer Prevention. 2012;13(7):3443–9.

24. Parsa P, Kandiah M, Mohd Zulkefli NA, Rahman HA. Knowledge and behavior regarding breast cancer screening among female teachers in Selangor, Malaysia. Asian Pacific Journal Of Cancer Prevention: APJCP. 2008;9(2):221–7.

25. Shah NM, Nan BLT, Hui NY, Islahudin FH, Hatah EM. Knowledge and perception of breast cancer and its treatment among Malaysian women: Role of religion. Tropical Journal of Pharmaceutical Research. 2017;16(4):955–62.

26. Al-Naggar RA, Bobryshev YV. Practice and Barriers of Mammography among Malaysian Women in the General Population. Asian Pacific Journal of Cancer Prevention. 2012;13(8):3595–600.

27. Hassan N, Ho WK, Mariapun S, Teo SH. A cross sectional study on the motivators for Asian women to attend opportunistic mammography screening in a private hospital in Malaysia: the MyMammo study. BMC Public Health. 2015;15:548.

28. Harmy MY, Norwati D, Noor NM, Amry AR. Knowledge and Attitude of Colorectal Cancer Screening Among Moderate Risk Patients in West Malaysia. Asian Pacific Journal of Cancer Prevention. 2011;12(8):1957–60.

29. Yusoff HM, Daud N, Noor NM, Rahim AA. Participation and Barriers to Colorectal Cancer Screening in Malaysia. Asian Pacific Journal of Cancer Prevention. 2012;13(8):3983–7.

30. Mohd Suan MA, Mohammed NS, Abu Hassan MR. Colorectal Cancer Awareness and Screening Preference: A Survey during the Malaysian World Digestive Day Campaign. Asian Pacific Journal Of Cancer Prevention: APJCP. 2015;16(18):8345–9.

31. Tin Tin S, Jun Yan G, Tan J, Abdul Rahim M, Yoganathan P, Nasirin Sallamun K, et al. Level of colorectal cancer awareness: a cross sectional exploratory study among multi-ethnic rural population in Malaysia. BMC Cancer. 2013;13(1):1–8.

32. Yan P, Yu CJ, Haris AAH, Yuan AS. Assessment of the level of knowledge of colorectal cancer among public at outpatient clinics in serdang hospital: A survey based study. Medical Journal of Malaysia. 2017;72(6):338–44.

33. Hashim SM, Fah TS, Omar K, Rashid MRA, Shah SA, Sagap I. Knowledge of Colorectal Cancer among Patients Presenting with Rectal Bleeding and its Association with Delay in Seeking Medical Advice. Asian Pacific Journal of Cancer Prevention. 2011;12(8):2007–11.

34. Al-Naggar RA, Al-Kubaisy W, Yap BW, Bobryshev YV, Osman MT. Attitudes towards colorectal cancer (CRC) and CRC screening tests among elderly Malay patients. Asian Pacific Journal Of Cancer Prevention: APJCP. 2015;16(2):667–74.

35. Naing C, Jun YK, Yee WM, Waqiyuddin SB, Lui LC, Shaung OY, et al. Willingness to take a screening test for colorectal cancer: A community-based survey in Malaysia. European Journal of Cancer Prevention. 2014;23(2):71–5.

36. Hilmi I, Hartono JL, Goh KL. Negative Perception in Those at Highest Risk - Potential Challenges in Colorectal Cancer Screening in an Urban Asian Population. Asian Pacific Journal of Cancer Prevention. 2010;11(3):815–22.

37. Akhtari-Zavare M, Latiff LA, Juni MH, Said SM, Ismail IZ. Knowledge of Female Undergraduate Students on Breast Cancer and Breast Self-examination in Klang Valley, Malaysia. Asian Pacific Journal Of Cancer Prevention: APJCP. 2015;16(15):6231–5.

38. Hadi MA, Hassali MA, Shafie AA, Awaisu A. Evaluation of breast cancer awareness among female university students in Malaysia. Pharmacy Practice (1886-3655). 2010;8(1):29–34.

39. Dinegde NG, Xuying L. Awareness of Breast Cancer among Female Care Givers in Tertiary Cancer Hospital, China. Asian Pacific journal of cancer prevention : APJCP. 2017;18(7):1977–83.

40. Tazhibi M, Feizi A. Awareness Levels about Breast Cancer Risk Factors, Early Warning Signs, and Screening and Therapeutic Approaches among Iranian Adult Women: A large Population Based Study Using Latent Class Analysis. BioMed Research International. 2014;2014:9.

41. Farid ND, Aziz NA, Al-Sadat N, Jamaludin M, Dahlui M. Clinical breast examination as the recommended breast cancer screening modality in a rural community in Malaysia; what are the factors that could enhance its uptake? PLoS One. 2014;9(9):e106469.

42. Oluwatosin OA, Oladepo O. Knowledge of breast cancer and its early detection measures among rural women in Akinyele Local Government Area, Ibadan, Nigeria. BMC Cancer. 2006;6(1):271.

43. Grunfeld EA, Ramirez AJ, Hunter MS, Richards MA. Women’s knowledge and beliefs regarding breast cancer. Br J Cancer. 2002;86(9):1373–8.

44. Junaibi RM, Khan SA. Knowledge and awareness of breast cancer among university female students in muscat, sultanate of oman- A pilot study. Journal of Applied Pharmaceutical Science. 2011;1:146–9.

45. Wood MF, Vial MC, Martinez-Gutierrez J, Mason MJ, Puschel K. Examining barriers for mammography screening compliance within a private hospital and an underserved primary care clinic in Santiago, Chile. Journal of the American College of Radiology : JACR. 2013;10(12):966–71.

46. Chua MS, Mok TS, Kwan WH, Yeo W, Zee B. Knowledge, perceptions, and attitudes of Hong Kong Chinese women on screening mammography and early breast cancer management. Breast J. 2005;11(1):52–6.

47. Nazzal Z, Sholi H, Sholi SB, Sholi MB, Lahaseh R. Motivators and barriers to mammography screening uptake by female health-care workers in primary health-care centres: a cross-sectional study. Lancet (London, England). 2018;391 Suppl 2:S51.

48. Wu T-Y, Hsieh H-F, West BT. Demographics and Perceptions of Barriers Toward Breast Cancer Screening Among Asian-American Women. Women & health. 2008;48(3):261–81.

49. Chua MS-T, Mok TSK, Kwan WH, Yeo W, Zee B. Knowledge, Perceptions, and Attitudes of Hong Kong Chinese Women on Screening Mammography and Early Breast Cancer Management. The Breast Journal. 2005;11(1):52–6.

50. Garbers S, Jessop DJ, Foti H, Uribelarrea M, Chiasson MA. Barriers to breast cancer screening for low-income Mexican and Dominican women in New York City. J Urban Health. 2003;80(1):81–91.

51. Baron-Epel O, Granot M, Badarna S, Avrami S. Perceptions of breast cancer among Arab Israeli women. Women & health. 2004;40(2):101–16.

52. Azaiza F, Cohen M. Health beliefs and rates of breast cancer screening among Arab women. Journal of women’s health (2002). 2006;15(5):520–30.

53. Wu TY, West B, Chen YW, Hergert C. Health beliefs and practices related to breast cancer screening in Filipino, Chinese and Asian-Indian women. Cancer detection and prevention. 2006;30(1):58–66.

54. Feldstein AC, Perrin N, Rosales AG, Schneider J, Rix MM, Glasgow RE. Patient Barriers to Mammography Identified During a Reminder Program. Journal of women’s health (2002). 2011;20(3):421–8.

55. Miller D, Livingstone V, Herbison P. Interventions for relieving the pain and discomfort of screening mammography. The Cochrane database of systematic reviews. 2008(1):Cd002942.

56. McCaffery K, Wardle J, Waller Jo. Knowledge, attitudes, and behavioral intentions in relation to the early detection of colorectal cancer in the United Kingdom. Preventive Medicine. 2003;36(5):525–35.

57. Power E, Simon A, Juszczyk D, Hiom S, Wardle J. Assessing awareness of colorectal cancer symptoms: Measure development and results from a population survey in the UK. BMC Cancer. 2011;11(1):366.

58. Yardley C, Glover C, Allen-Mersh TG. Demographic factors associated with knowledge of colorectal cancer symptoms in a UK population-based survey. Ann R Coll Surg Engl. 2000;82(3):205–9.

59. McVeigh TP, Lowery AJ, Waldron RM, Mahmood A, Barry K. Assessing awareness of colorectal cancer symptoms and screening in a peripheral colorectal surgical unit: a survey based study. BMC Surg. 2013;13:20-.

60. Nasaif HA, Al Qallaf SM. Knowledge of Colorectal Cancer Symptoms and Risk Factors in the Kingdom of Bahrain: a Cross-Sectional Study. Asian Pac J Cancer Prev. 2018;19(8):2299–304.

61. Jalleh G, Donovan RJ, Lin C, Slevin T, Clayforth C, Pratt IS, et al. Beliefs about bowel cancer among the target group for the National Bowel Cancer Screening Program in Australia. Australian and New Zealand Journal of Public Health. 2010;34(2):187–92.

62. Christou A, Thompson SC. Colorectal cancer screening knowledge, attitudes and behavioural intention among Indigenous Western Australians. BMC Public Health. 2012;12(1):528.

63. Saeed RS, Bakir YY, Alkhalifah KH, Ali LM. Knowledge and Awareness of Colorectal Cancer among General Public of Kuwait. Asian Pacific journal of cancer prevention : APJCP. 2018;19(9):2455–60.

64. Muthukrishnan M, Arnold LD, James AS. Patients’ self-reported barriers to colon cancer screening in federally qualified health center settings. Prev Med Rep. 2019;15:100896-.

65. Tessaro I, Mangone C, Parkar I, Pawar V. Knowledge, barriers, and predictors of colorectal cancer screening in an Appalachian church population. Preventing chronic disease. 2006;3(4):A123.

66. Bo ZM, Ghevariya V, Ahluwalia M, Veerabhadrappa K, Villani GM, Anand S. Barriers to colorectal cancer screening among African American population. Journal of Clinical Oncology. 2010;28(15_suppl):1568-.

67. Wong MCS, Ching JYL, Hirai HH, Lam TYT, Griffiths SM, Chan FKL, et al. Perceived Obstacles of Colorectal Cancer Screening and Their Associated Factors among 10,078 Chinese Participants. PLOS ONE. 2013;8(7):e70209.

68. Stacy R, Torrence WA, Mitchell CR. Perceptions of Knowledge, Beliefs, and Barriers to Colorectal Cancer Screening. Journal of Cancer Education. 2008;23(4):238–40.

69. Leung DYP, Chow TT, Wong EML. Cancer-Related Information Seeking and Scanning Behaviors among Older Chinese Adults: Examining the Roles of Fatalistic Beliefs and Fear. Geriatrics (Basel). 2017;2(4):38.

70. Guessous I, Dash C, Lapin P, Doroshenk M, Smith RA, Klabunde CN. Colorectal cancer screening barriers and facilitators in older persons. Preventive Medicine. 2010;50(1):3–10.

71. Lee M-S, Azmiyaty Amar Ma’ Ruf C, Nadhirah Izhar DP, Nafisah Ishak S, Wan Jamaluddin WS, Ya’acob SNM, et al. Awareness on breast cancer screening in Malaysia: a cross sectional study. Biomedicine (Taipei). 2019;9(3):18-.

72. Schliemann D, Paramasivam D, Dahlui M, Cardwell CR, Somasundaram S, Ibrahim Tamin Nsb, et al. Change in public awareness of colorectal cancer symptoms following the Be Cancer Alert Campaign in the multi-ethnic population of Malaysia. BMC Cancer. 2020;20(1):252.

73. Sindhu CK, Nijar AK, Leong PY, Li ZQ, Hong CY, Malar L, et al. Awareness of Colorectal Cancer among the Urban Population in the Klang Valley. Malays Fam Physician. 2019;14(3):18–27.

